# AI-Driven Plasma Denaturation Profiling for Multi-Cancer Detection

**DOI:** 10.1101/2025.03.26.25324680

**Authors:** Philipp O. Tsvetkov, Rémi Eyraud, Stéphane Ayache, Viktoriia E. Baksheeva, Alexandre Bertucci, Alice Mogenet, Pascale Tomasini, Bernadette De Rauglaudre, Laetitia Dahan, Caroline Gaudy-Marquestre, Shanmugha Sri Siva Kalidindi, Christophe Buffat, Caroline Dehais, Alexandre Astier, l’Houcine Ouafik, Svetlana Gorkhova, Emeline Tabouret, François Devred

## Abstract

Many cancers cannot be detected early due to lack of effective disease biomarkers, leading to poor prognosis. We applied an existing biophysical technology nanoDSF in a novel way to answer this unmet biomedical need. We developed a breakthrough digital biomarker method for cancer detection based on AI-classification of plasma denaturation profiles (PDPs) obtained by nanoDSF technology. PDPs from 300 plasma samples from patients with melanoma, brain, digestive or lung cancers were automatically distinguished from ‘healthy’ profiles with an accuracy of 94%. Moreover, our method was able to distinguish different types of cancers from each other with an accuracy of 80%, making it an effective way to help cancer diagnosis and monitoring. Our technology thus paves the way for a long-sought multi-cancer early detection (MCED) test that is blood-based, cost-effective and easy-to-implement in any clinical setting.

## Main text

Prognosis and survival of cancer patients is much worse if the disease is not detected early. However, efficient and non-invasive ways to detect cancer at early stages are not available for most types of cancer. Significant efforts have been concentrated on searching for molecular biomarkers that can be used for cancer detection. However, the field is beginning to realize that single molecular biomarkers cannot capture the underlying complexity of cancer nor can they be used alone to establish an early and precise diagnosis. Rather than looking for individual biomarkers for each cancer, many teams are now developing blood-based multi-cancer early detection (MCED) approaches based on analyzing multimodal signals in an integrated manner, enabled by the seemingly unlimited possibilities offered by AI ^1–5^. One of these approaches is our recently developed cancer detection method that is based on plasma denaturation profiling by nanoDSF ^6,7^. This method relies on detecting changes in the thermal stability of circulating proteins resulting from the tumor presence in the organism and leading to specific plasma denaturation profiles (PDPs) ^8,9^. AI-powered analysis of these profiles allowed us to efficiently distinguish profiles of patients with glioma from that of healthy individuals ^6^ and even discriminate between molecular subtypes of glioma ^7^.

We are now extending this approach to other cancer types using a novel AI-nanoDSF workflow and plasma samples from patients with melanoma, digestive adenocarcinoma, brain or lung cancers before the initiation of a new treatment line (Figure 1A-C). Using a nanoDSF instrument we obtained PDPs, consisting of the following eight outputs over temperature for a given plasma sample: F_330_, F_350_, F_330_/F_350_, A_350_ as well as their derivatives (see methods for details). These PDPs were used as inputs for five machine learning algorithms (Logistic Regression (LR), Support Vector Machines (SVM), Random Forest (RF), Extreme Gradient Boosting (XGBoost), and Deep Neural Networks (Deep)) before applying the Bagging Method that does the weighted average of the output of these five algorithms to increase robustness ^10^. For each algorithm we used 80% of the data (training set) to search for the best combination of hyperparameters using a 10-fold cross-validation grid search. We then trained a model using the found hyperparameter combination, and evaluated the learned model on the remaining 20% of the data. To ensure the maximal robustness of the results, we repeated this protocol 5 times using different evaluation and training sets. All reported results represent the average over these 5 experimental runs (see Methods for details).

**Figure 1.**
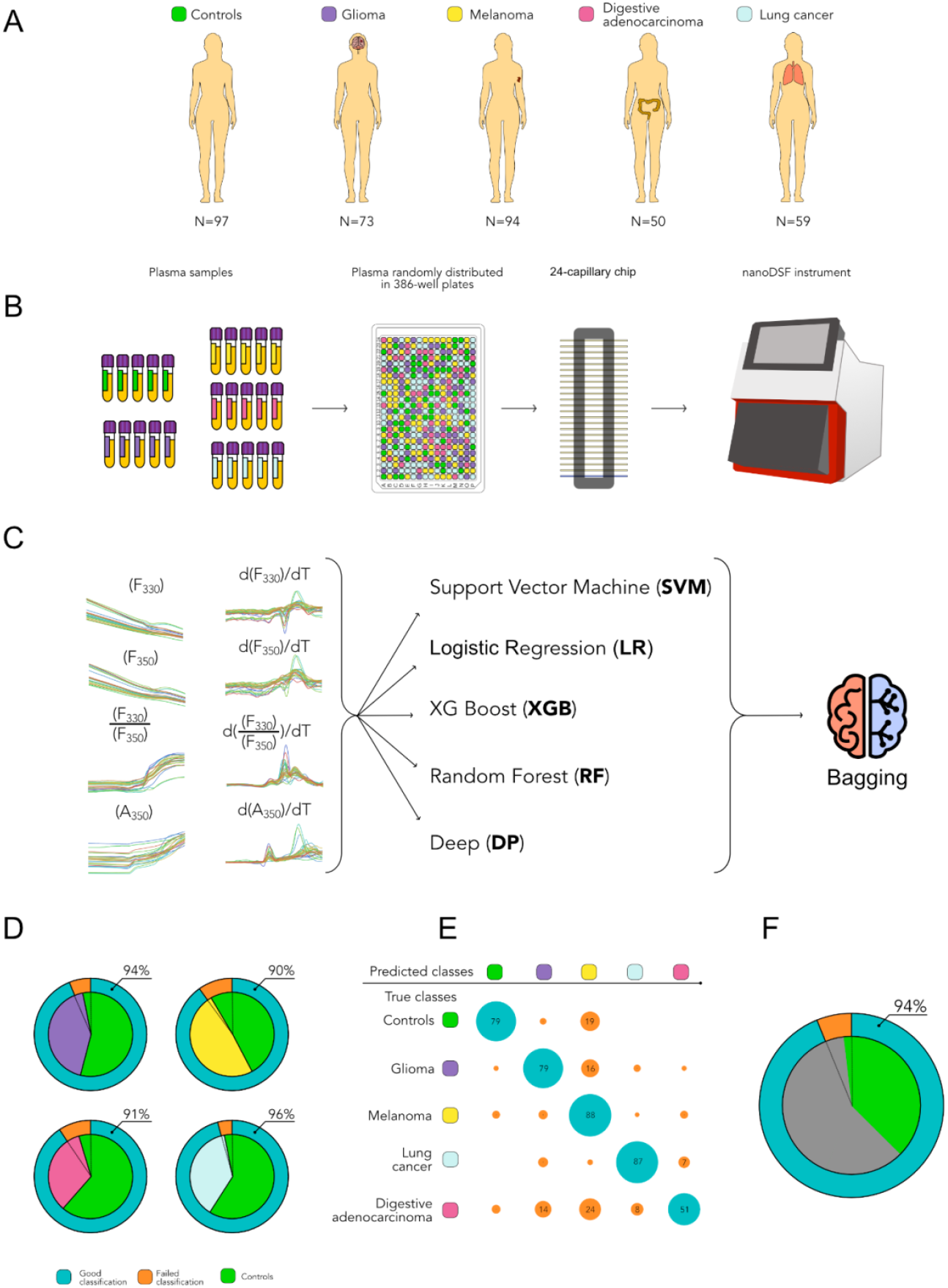
A: Five cohorts used in the study. B: Workflow for Obtaining PDPs from Plasma Samples Using NanoDSF. C: PDPs from NanoDSF instrument used as input data for AI algorithms. D: Assay 1 charts showing accuracy, and the rates of successful (blue) and false (orange) classifications of PDPs separately across four cancers (glioma shown in violet, melanoma in yellow, digestive cancer in pink, and lung cancer in light blue) and healthy controls (in green). E: Assay 2 confusion matrix of *Bagging* model. Numerical values show the corresponding percentage of a given class classified as such, size of a circle its relative value; blue circles correspond to correct classification and orange circles to misclassification. The overall average accuracy of this model over all the classes is 80±6 %. F: Assay 3 chart showing accuracy, and the rates of successful and failed classifications of PDPs of four combined cancer (gray) and healthy controls (green).

We first applied this new AI workflow to verify that each cancer tested induces specific PDPs (Assay 1). Using 276 plasma samples from patients with brain cancer, melanoma, digestive adenocarcinoma, or lung cancer (Supplementary Table 1), we trained algorithms to distinguish PDPs of each cancer patient group from that of healthy individuals used as controls. Our workflow allowed us to distinguish each cancer from the controls with an accuracy of 96.1% for lung cancer, 93.9 % for glioma, 91.7% for digestive adenocarcinoma and 90% for melanoma (Figure 1D, Table 1). The detailed results of all algorithms are shown in the Supplementary Tables 2.1-2.4. These results indicate that PDPs constitute a digital biomarker for each cancer tested and could be used to differentiate the cancer patient’s plasma, from that of controls. If a patient’s symptoms or a radiological lesion orient towards a potential cancer diagnosis, this simple blood-based approach could be used to support or rule out the diagnosis of cancer.

**Table 1.**

|  | Glioma* | Melanoma** | Lung*** | Digestive**** | Cancer |
| --- | --- | --- | --- | --- | --- |
| Accuracy % | 94±4 | 90±5 | 96±4 | 91±4 | 94±3 |
| False positives | 1.0±0.8 | 4±3 | 1±2 | 1±1 | 3±2 |
| False negativ | 1.0±0.8 | 0.2±0.5 | 0.4±0.6 | 1.0±0.8 | 1.6±0.9 |
| Sensitivity | 0.93 | 0.99 | 0.97 | 0.90 | 0.97 |
| Specificity | 0.95 | 0.81 | 0.96 | 0.93 | 0.84 |
| Precision | 0.93 | 0.84 | 0.94 | 0.87 | 0.95 |

The outputs of the Bagging method for each cancer types. The accuracy values (+/- standard deviation) correspond to the average over 5 repetitions of the evaluation protocol (see Methods for details). Number of false positives and negatives are the overall average over all the 5 runs. The other metrics are calculated from these latter values. Number of samples in each of the 5 tests: 19-20 controls (out of 97 total), 14-15 glioma (out of 73 total), 18-19 melanoma (out of 94 total), 11-12 lung cancer (out of 59 total), 10 digestive adenocarcinoma (out of 50 total)

We then set out to develop a test that would determine the subtype of cancer when a malignant process is suspected but the primary tumor remains undetermined (Assay 2). We therefore trained the models to classify five different groups of samples concomitantly - the four cancer tested and controls. The following important observations can be made from the confusion matrix of the Bagging Model (Figure 1E, blue circles corresponding to the percentage of correctly classified samples). First, very few cancer samples were misclassified as controls : 1% for glioma, 3% for melanoma, 0% for lung and 4% for digestive adenocarcinoma (see first column). Second, pulmonary cancer and melanoma had the highest numbers of correctly classified cancer samples (87 and 88% respectively), though the detection of pulmonary cancer was more specific (see 4th column). Third, the model performed less well for the digestive adenocarcinoma group (see 5th line), likely explained by its heterogeneity (it gathers colo-rectal and pancreatic cancers). Overall, the accuracy for this multiclass analysis (correct classifications out of total number of classifications) was 80% (+/−6), showing that different types of cancers can be distinguished from one another by their PDPs. This approach which uses PDPs as digital biomarkers can thus be used to develop a multi-cancer test that will contribute to diagnosis when one of the given cancers is suspected.

Finally, we set out to test if our approach also can be used to identify a general cancer signature, independently of its tissue origin (Assay 3). We used an extended cohort of 300 cancer patients (supplementary table 3) to train and test the algorithms. We were able to distinguish cancers from controls with an accuracy of 94% (Figure 1F, Table 1, supplementary table 3). Thus, this approach could potentially be used as a screening test to detect cancer of any origin, especially if a clinical or radiological symptom points toward a cancer diagnosis, to save time and improve patient prognosis.

Altogether, these three assays constitute a test that answers a major unmet need in oncology. Fast detection of cancer is a major public health goal, as rapid diagnosis determines prognosis in the majority of cases. However, apart from specific indications in certain countries, there is no mass screening for cancers due to lack of a reliable, reproducible, minimally invasive and inexpensive test. Similarity to other liquid biopsy based methods ^1,3^, our test is designed to address this important issue. However, our test is particularly easy to implement on a large-scale basis and offers significant advantages over all other competitive methods: (i) Our test requires a very small amount of a standard plasma preparation used for a routine blood-work. (ii) Sample analysis can be automated and performed in less than one hour, allowing high throughput analysis of numerous samples. (iii) No special analysis skills are required to interpret the output of the instrument, as data analysis is completely automated by incorporated AI algorithms. (iv) The instrument is much less expensive compared to that used by other methods, such as methylation profile sequencers or tumor cell / exosome extractors. (v) At last, this test does not require potentially expensive and perishable reagents. Thus our novel test has all the key requirements to be used as a large-scale multi-cancer screening test to promote fast and cost-effective cancer detection in the general population.

Our test can also be implemented to manage the cancer patients in a hospital setting where oncologists are often confronted with the discovery of secondary lymph node or metastatic localizations. The primary cause of these lesions is often known until several days or even weeks later, delaying effective patient management. Our test could rapidly indicate the tissue of cancer origin, thus having a significant impact on patient prognosis, especially for solid tumors. Indeed, unlike in hematology where liquid biopsies are genuine surrogate markers for diagnosis and monitoring of the disease, molecular biomarkers and circulating tumor cell analysis are much less established in solid oncology ^11,12^.

In conclusion, using PDPs as digital biomarkers constitute a major breakthrough in oncology as it allows a single, cost-effective, and minimally invasive assay to detect various cancers across different organs without previous knowledge of molecular biomarkers. Extending this approach to many other cancers will be needed in the future to establish a more complete multi-cancer early detection test.

## Data Availability

All data produced in the present study are available upon reasonable request to the authors

## Acknowledgments

This study was partly supported by research funding from the Cancéropôle PACA, Institut National du Cancer and Région Sud, MIC grant from ITMO Cancer of Aviesan, Patient association ARTC Sud and by INCa-DGOS-Inserm_12560 grant (SiRIC CURAMUS). We would also like to thank AP-HM Tumor Bank (authorization number CRB BB-0033-00097) and the Onconeurotek Tumor Bank (APHP).

## METHODS

### Cohorts

Patient cohort (Supplementary Table 1) consisted of 300 patients included at Assistance Publique Hôpitaux de Marseille (APHM) from January 2009 to May 2020 and 33 patients included at La Pitié Salpétrière Hospital (Paris) from November 2008 to September 2016. Eligible patients included were those aged 16 years or older with melanoma, digestive cancers, lung cancers or glioma for whom plasma samples were available at the time of diagnosis or recurrence, before the initiation of new treatment line. Clinical, histomolecular and radiological data were recorded. All patients provided written informed consent in accordance with institutional, national guidelines and the Declaration of Helsinki (PADS 22-98). 97 plasma samples from healthy adult volunteers were used as controls.

### Plasma samples

Blood samples were collected into EDTA tubes, separated by centrifugation (2000g, 10 minutes, 20°C, twice) within 30 minutes and then stored at − 80°C. No other specific purification step was added in order not to perturb the interactome or alter the chemical state of plasma proteins. Before nanoDSF analysis, samples were thawed rapidly at 37°C.

### Plasma denaturation profile

Plasma samples were loaded to 10 μL standard 24-capillary chips from Nanotemper and then scanned using nanoDSF Prometheus NT.Plex 24 instrument equipped with scattering module (Nanotemper) at 5% of laser power and 1°C/min heating rate in order to obtain the denaturation profiles in the range from 15 to 95°C. Raw data were exported into datasets that contained all of the instrument outputs: fluorescence at 330 and 350 nm (F_330_ and F_350_), the ratio of these values (F_330_/F_350_) as well as absorbance at 350 nm (A_350_). The plasma denaturation profiles (PDPs) are defined as the set of the 8 outputs from Prometheus NT.Plex for a given plasma sample, here the 4 curves F_330_, F_350_, F_330_/F_350_, A_350_ and their derivatives.

### AI evaluating protocol

To evaluate the quality of our approach on unseen data, we first splitted the PDPs samples into two sets, 20% of them for evaluation and 80% for training, in a randomized stratified manor: the proportion of data of each type has to be the same in the 2 sets than in the whole set of data. All used learning algorithms come with hyperparameters: these algorithms are functions whose output is determined by different values that need to be fixed beforehand. Since the chosen values have a huge impact on the quality of the model that the algorithm learns from the training data, it is extremely important to find the combination of values that will allow the algorithm to output the best model possible. To do so, we trained the algorithm with each potential combination and evaluated its outputted model, a process called grid search ^13^. Given one combination, we ran a stratified 10-fold cross-validation on the training set: data were randomly separated into 10 stratified samples of the same size. The under investigation algorithm was then trained on the 9 samples and the model it outputs was tested on the 10th. The procedure was repeated 10 times so that each fold of data is used once as a test sample. For the Deep approach, performing cross-validation to select optimal hyperparameters being too costly due to higher number of hyperparameters and longer training time, we grid-searched a total of 24 combinations of hyperparameters listed in Supplementary Table 5. We then computed the average evaluation metric on the set left out and assigned this score to this combination of hyperparameters. This approach tackles the issue of statistical instability and experimental artifacts ^14^.

Once we evaluated the metric - here the accuracy - for all combinations of hyperparameters, we selected the best one. Using these values, we then ran the learning algorithm on the whole training set to obtain a trained model, whose capability was then evaluated on the 20% of data left apart from the initial split. As this 20/80 initial split may have an impact on the evaluation of the quality of the approach, we ran this protocol with 5 different splits and computed the average evaluation metric and its deviation, which are the values reported throughout this paper.

### AI processing : Algorithm implementation

The code used was written in Python (version 3.8.10). The data preparation was done using the pandas library (version 1.4.0) while the machine learning algorithms were run using the scikit-learn toolbox (version 1.1.0), except for the Deep one which used Tensorflow (version 2.11). PDPs obtained from the nanoDSF instrument were interpolated using InterpolatedUnivariateSpline from the scipy.interpolate (version 1.5.4) module in order to ensure the same temperature alignment and the same dimension (1,200 for each curve) for all data.

The data were first split into a train set and a test set: using the train_test_split function from sklearn.model_selection module with the parameter test_size set to 0.2 and the other parameters set to default. We then created pipelines using the method Pipeline from the module sklearn.pipeline with the function StandardScaler from the sklearn.preprocessing module as first parameter and the corresponding learning algorithm as a second parameter. This ensured homogeneous et relevant use of this normalization throughout the experiments.

The research of the best combination of hyperparameters was conducted using the function GridSearchCV from the module sklearn.model_selection with the following parameter settings: estimator set to the previously created pipeline, param_grid set to the values described in supplementary table 5, scoring set to ‘accuracy’, and cv set to the output of the function StratifiedKFold from sklearn.model_selection with n_splits=10, shuffle=True, random_state=42. The different tested implementations are: (1) LogisticRegression from the linear_model module; (2) SVC from the svm module; (3) RandomForestClassifier from the ensemble module; (4) XGBClassifier from the module xgboost. (5) Deep convolutional network using Tensorflow/Keras (see supplementary methods)

## Notes

### Competing Interest Statement

The authors have declared no competing interest.

### Funding Statement

This study was partly funded by Canceropole PACA, Institut National du Cancer and Region Sud, MIC grant from ITMO Cancer of Aviesan, Patient association ARTC Sud and by INCa-DGOS-Inserm_12560 grant (SiRIC CURAMUS).

### Author Declarations

Ethic committee of Assistance Publique des Hopitaux de Marseille AP-HM (CRB BB-0033-00097) gave ethical approval for this work.

