## supplementary data for "AI-Driven Plasma Denaturation Profiling for Multi-Cancer Detection"

Supplementary Table 1 - Patient characteristics.

| **Factors** | **Melanoma, N=94** | **Digestive adenocarcinoma (colo-rectal / pancreatic cancer), N=50** | **Lung cancers, N=59** | **Glioma, N=73** |
|---|---|---|---|---|
| Age (median, range) | 64.2 (16.2-89.7) | 66.5 (33.6-8) | 68.0 (33.2-90.5) | 60.1 (21.5-80.9) |
| Gender (W/M) | 36/58 (62/38) | 20/30 (40/60) | 28/31 (48/52) | 29/44 (40/60) |
| OMS | | | | |
| 0 | 86 (93) | 14 (28) | 29 (49) | |
| 1 | 7 (6) | 22 (44) | 20 (34) | |
| 2 | | 7 (14) | 8 (14) | |
| ≥ 3 | | 1 (2) | 2 (3) | |
| NA | 1 (1) | 6 (12) | | |
| KPS (median, range) | | | | 80 (40-100) |
| Metastases | | | | |
| Yes | 3 (3) | 34 (68) | 45 (76) | |
| No | 90 (96) | 12 (24) | 14 (24) | |
| NA | 1 (1) | 4 (8) | | |
| Surgical resection | | | | |
| Yes | | 26 (52) | 8 (14) | 55 (75) |
| No | | 23 (46) | 51 (86) | 15 (21) |
| NA | | 1 (2) | | 3 (4) |
| Systemic treatments following plasma collection | | | | |
| Yes | 11 (12) | 46 (92) | 51 (86) | 63 (86) |
| No | 82 (88) | 0 | 8 (14) | 10 (14) |
| NA | 1 (1) | 4 (8) | | |
| Radiotherapy | | | | |
| Yes | | | | 52 (71) |
| No | | | | 21 (29) |
| Molecular alterations | | | | |
| *BRAF* V600E mutation | 15 (16) | | | |
| *EGFR* mutation | | | 18 (30) | |
| Smoker | | | 37 (63) | |

### Supplementary Table 2.1. Glioma vs. control (Assay 1)

| | LR | RF | SVM | XGBoost | Deep | Bagging |
|---|---|---|---|---|---|---|
| accuracy | 93±5 | 92±3 | 94±6 | 88±10 | 88±3 | 94±4 |
| False + | 1.0±0.7 | 1.6±1.3 | 1.2±1.1 | 2.2±1.6 | 2.4±0.5 | 1±0.71 |
| Fakse - | 1.4±1.1 | 1.0±0.7 | 0.8±0.8 | 2.2±1.6 | 1.6±0.9 | 1±0.71 |
| Sensitivity | 0.907 | 0.933 | 0.947 | 0.853 | 0.893 | 0.933 |
| Specificity | 0.944 | 0.911 | 0.933 | 0.878 | 0.867 | 0.947 |
| Precision | 0.932 | 0.897 | 0.922 | 0.853 | 0.848 | 0.933 |

The accuracy values (± standard deviation) correspond to the average over 5 repetitions of the evaluation protocol (see Methods for details). Number of false positives and negatives are the overall average over all the 5 runs. Sensitivity, Specificity and precision are calculated from these values. Number of samples in each of the 5 tests: 19-20 controls, 14-15 glioma, overall number of samples: 97 controls, 73 glioma.

### Supplementary Table 2.2. Melanoma vs. control (Assay 1)

| | LR | RF | SVM | XGBoost | Deep | Bagging |
|---|---|---|---|---|---|---|
| accuracy | 87±7 | 87±6 | 86±6 | 87±4 | 89±3 | 90±6 |
| False + | 4.2±2.6 | 4.0±1.2 | 4.0±2.7 | 4.0±1.9 | 3.4±1.8 | 3.6±2.07 |
| False - | 0.6±0.9 | 0.8±0.8 | 1.4±1.1 | 0.8±0.9 | 0.8±1.1 | 0.2±0.45 |
| Sensitivity | 0.968 | 0.958 | 0.926 | 0.958 | 0.958 | 0.989 |
| Specificity | 0.779 | 0.789 | 0.789 | 0.789 | 0.821 | 0.811 |
| Precision | 0.814 | 0.820 | 0.815 | 0.820 | 0.843 | 0.840 |

The accuracy values (± standard deviation) correspond to the average over 5 repetitions of the evaluation protocol (see Methods for details). Number of false positives and negatives are the overall average over all the 5 runs. Sensitivity, Specificity and precision are calculated from these values. Number of samples in each of the 5 tests: 19-20 controls, 18-19 melanoma, overall number of samples: 97 controls, 94 melanoma.

### Supplementary Table 2.3. Lung cancer vs. control (Assay 1)

| | LR | RF | SVM | XGBoost | Deep | Bagging |
|---|---|---|---|---|---|---|
| accuracy | 92±5 | 96±4 | 91±5 | 94±5 | 95±2 | 96±4 |
| False + | 1.6±1.5 | 0.0±0.0 | 2.0±1.0 | 0.2±0.4 | 1.0±0.0 | 0.8±1.30 |
| False - | 0.8±0.8 | 1.2±1.1 | 0.8±0.8 | 1.6±1.1 | 0.4±0.5 | 0.4±0.55 |
| Sensitivity | 0.926 | 0.900 | 0.933 | 0.867 | 0.967 | 0.967 |
| Specificity | 0.789 | 1.0 | 0.895 | 0.989 | 0.947 | 0.958 |
| Precision | 0.815 | 1.0 | 0.848 | 0.981 | 0.921 | 0.935 |

The accuracy values (± standard deviation) correspond to the average over 5 repetitions of the evaluation protocol (see Methods for details). Number of false positives and negatives are the overall average over all the 5 runs. Sensitivity, Specificity and precision are calculated from these values. Number of samples in each of the 5 tests: 19-20 controls, 11-12 lung cancer, overall number of samples: 97 controls, 59 lung cancer.

### Supplementary Table 2.4. Digestive adenocarcinoma vs. control (Assay 1)

Test: 19 controls, 10 Digestive Adenocarcinoma

| | LR | RF | SVM | XGBoost | Deep | Bagging |
|---|---|---|---|---|---|---|
| accuracy | 87±7 | 90±5 | 86±8 | 91±2 | 90±4 | 93±1 |
| False + | 2.6±2.1 | 1.4±1.1 | 1.8±0.8 | 2.0±0.7 | 2.4±0.5 | 1.4±0.89 |
| False - | 1.2±1/1 | 1.6±1.3 | 1.6±1.1 | 0.6±0.5 | 0.4±0.5 | 1±0.71 |
| Sensitivity | 0.880 | 0.840 | 0.840 | 0.940 | 0.960 | 0.900 |
| Specificity | 0.863 | 0.926 | 0.905 | 0.895 | 0.874 | 0.926 |
| Precision | 0.772 | 0.857 | 0.824 | 0.825 | 0.800 | 0.865 |

The accuracy values (± standard deviation) correspond to the average over 5 repetitions of the evaluation protocol (see Methods for details). Number of false positives and negatives are the overall average over all the 5 runs. Sensitivity, Specificity and precision are calculated from these values. Number of samples in each of the 5 tests: 19-20 controls, 10 digestive adenocarcinoma, overall number of samples: 97 controls, 50 digestive carcinoma.

Supplementary Table 3. Patient characteristics of the extended cohort.

| **Factors** | **All tumors, N=300** |
| --- | --- |
| Age | 63.8 |
| (median, range) | (16.2-95.5) |
| Gender (W/M) | 121/179 (40/60) |
| Lung | 59 (20) |
| ADK | 53 |
| Squamous cells | 4 |
| Other | 2 |
| Melanoma | 94 (31) |
| SSM | 61 |
| NM | 15 |
| AL | 2 |
| Dubreuilh | 1 |
| Other | 15 |
| Glioma | 73 (24) |
| Astrocytoma | 11 |
| Oligodendroglioma | 11 |
| Glioblastoma | 51 |
| Digestive tumors | 74 (25) |
| Esophagus | 5 |
| Cardia | 5 |
| Stomach | 6 |
| Anal | 3 |
| Cholangiocarcinoma | 5 |
| Pancreas | 23 |
| Rectal | 8 |
| Colon | 19 |

| | LR | SVM | RF | XGBoost | Deep | Bagging |
|---|---|---|---|---|---|---|
| Accuracy | 91±3 | 93±3 | 93±2 | 89±4 | 92±3 | 94±3 |
| False positives | 3.8 ±3.0 | 4.2±1.3 | 4.4±1.8 | 5.8±1.6 | 4.6±2.3 | 3±1.87 |
| False negatives | 3.4±1.7 | 1.2±1.8 | 1.0±0.7 | 3±1.2 | 1.2±0.8 | 1.6±0.89 |
| Sensitivity | 0.943 | 0.980 | 0.983 | 0.950 | 0.980 | 0.973 |
| Specificity | 0.800 | 0.779 | 0.768 | 0.695 | 0.758 | 0.842 |
| Precision | 0.937 | 0.933 | 0.931 | 0.908 | 0.927 | 0.951 |

Supplementary Table 4. All cancers vs controls (Assay 3).

The accuracy values correspond to the average over 5 repetitions of the evaluation protocol explained previously, standard deviation is reported next to symbol '±'. Number of false positives and negatives are the overall average over all the runs. The other metrics are calculated from these latter values.

Supplementary Table 5. Values tested for the hyperparameters of each learning algorithm.

| **Algorithm** | **Hyperparameters** | | | | | |
|---|---|---|---|---|---|---|
| **LR** | | penalty | C | | max_iter | |
| | **values** | {‘l1’, ‘l2’} | {100, 1000, 10000, 100000000} | | 100, 300, 500, 600, 900, 1000, 1200} | |
| **RF** | | min_samples_leaf | max_depth | min_samples_split | criterion | |
| | **values** | {1, 2, 3, 4, 5, 6} | {1, 2, 3, 4, 5, 6} | {2, 3, 4, 5, 6} | {'gini', 'entropy', 'log_loss'} | |
| **SVM** | | C | | kernel | | |
| | **values** | [100, 1000, 1000, 1000000000] | | {‘poly’: degree in {1, 2, 3}, ‘rbf’: gamma in {'scale', 'auto', 0.1, 0.5}} | | |
| **XGBoost** | | learnin_rate | max_depth | min_child_weight | subsample | n_estimators |
| | **values** | {.1, .2, .3} | {1, 2, 3, 4, 5, 6} | {1, 2, 3, 4, 5, 6} | {1, 0, 0.5, 0.1} | {100, 500, 1000} |
| **Deep** | | Jittering noise | Dropout | Weight decay | Base nb_filters | Learning_rate |
| | **values** | {.0001, .001} | {.1, .2} | {.01, .1} | {128, 256, 512} | .0001 |

Supplementary figure 1.

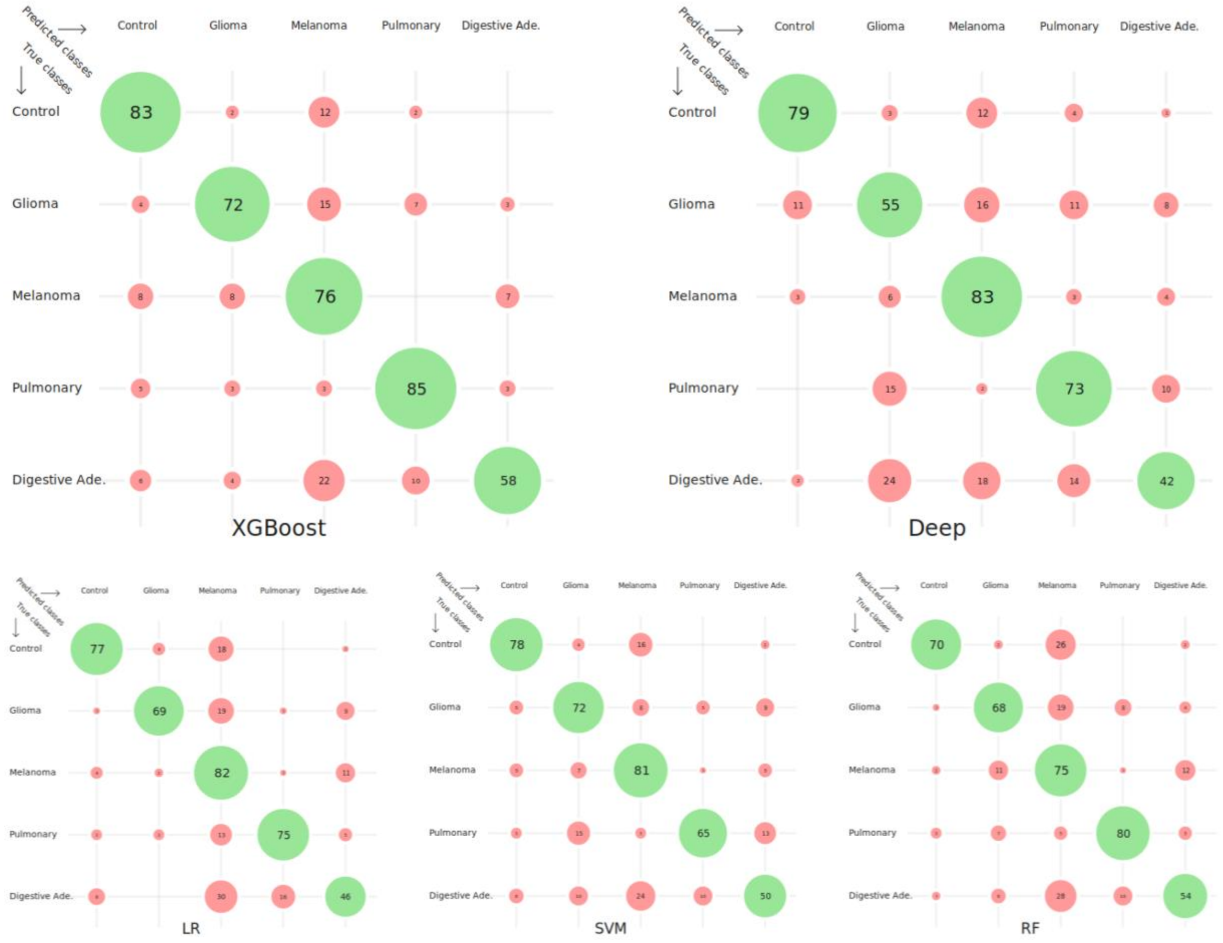

Figure S1. Confusion matrices comparing the performances of 5 classification algorithms for 5 classes predictions. Numerical values show the corresponding percentage, size of a circle its relative value, green circles correspond to correct classification. The overall average accuracy of LR over all the classes is 72 ± 4, of SVM is 70.27±4.17, of RF is 71.62±6.50, of XGBoost is 75.95±5.00 and of Deep is 69.19±5.60.

### Supplementary methods

(1) Logistic Regression (LR) is a binomial regression that provides the probability of unseen data belonging to a class. This algorithm comes with several hyperparameters: a constant C that sets the regularization strength, a weighting function for unbalanced classes, the maximum number of iterations, the penalty function used, the solver to be used.

(2) Random Forest (RF) is an ensemble method algorithm that learns several decision trees by sampling the data: its classification is made by a majority vote of all the learned trees. This algorithm comes with a large amount of hyperparameters: a weighting function for unbalanced classes, the criterion to create the trees, the tree maximal depth, a constant to decide when a node should be considered a leaf.

(3) Support Vector Machines (SVM) aims to find a hyperplane separating at best the classes. The power of SVM comes from their ability to project the data via a kernel in a different space where they are more likely to be linearly separable. This algorithm has several hyperparameters to be tuned : a constant C that sets the softness of the margin, the kernel used, and the parameters of each kernel.

(4) Extreme Gradient Boosting (XGBoost) is an ensemble method algorithm that is closely related to RF but with a different way to sample data for the learning of each tree. It has the advantage over other boosting algorithms, like AdaBoost, to scale well when the number of data increases. Its hyperparameters are the same as Random Forest.

(5) Deep Neural Networks (Deep) are usually adapted to cases where data are strongly structured and available in a larger quantity. Here we consider the PDPs data as time series that we normalized using Batch Normalization in input, followed by jittering data augmentation (adding gaussian noise). The rest of the architecture is a “small” ResNet1D composed of 6 convolutional layers with residual connections, dropout for regularization and a final dense layer as a linear classifier. The model has about 20M parameters, we trained it using Adam optimizer with weight decay regularization to avoid overfitting.
